# Risk Score Stratification of Alzheimer’s Disease and Mild Cognitive Impairment using Deep Learning

**DOI:** 10.1101/2020.11.09.20226746

**Authors:** Sanjay Nagaraj, Tim Q Duong

**Affiliations:** Department of Radiology, Renaissance School of Medicine, Stony Brook University, New York 11794

**Author notes:** **Funding** None. **Disclaimer** NA.

**Keywords:** Machine learning, artificial intelligence dementia, Alzheimer’s disease, mild cognitive impairment, Dementia, Deep Learning

## Abstract

Alzheimer Disease (AD) is a progressive neurodegenerative disease that can significantly impair cognition and memory. AD is the leading cause of dementia and affects one in ten people age 65 and older. Current diagnoses methods of AD heavily rely on the use of Magnetic Resonance Imaging (MRI) since non-imaging methods can vary widely leading to inaccurate diagnoses. Furthermore, recent research has revealed a substage of AD, Mild Cognitive Impairment (MCI), that is characterized by symptoms between normal cognition and dementia which makes it more prone to misdiagnosis.

A large battery of clinical variables are currently used to detect cognitive impairment and classify early mild cognitive impairment (EMCI), late mild cognitive impairment (LMCI), and AD from cognitive normal (CN) patients. The goal of this study was to derive a simplified risk-stratification algorithm for diagnosis and identify a few significant clinical variables that can accurately classify these four groups using an empirical deep learning approach. Over 100 variables that included neuropsychological/neurocognitive tests, demographics, genetic factors, and blood biomarkers were collected from EMCI, LMCI, AD, and CN patients from the Alzheimer’s Disease Neuroimaging Initiative (ADNI) database. Feature engineering was performed with 5 different methods and a neural network was trained on 90% of the data and tested on 10% using 10-fold cross validation. Prediction performance used area under the curve (AUC) of the receiver operating characteristic analysis.

The five different feature selection methods consistently yielded the top classifiers to be the Clinical Dementia Rating Scale - Sum of Boxes (CDRSB), Delayed total recall (LDELTOTAL), Modified Preclinical Alzheimer Cognitive Composite with Trails test (mPACCtrailsB), the Modified Preclinical Alzheimer Cognitive Composite with Digit test (mPACCdigit), and Mini-Mental State Examination (MMSE). The best classification model yielded an AUC of 0.984, and the simplified risk-stratification score yielded an AUC of 0.963 on the test dataset.

Our results show that this deep-learning algorithm and simplified risk score derived from our deep-learning algorithm accurately diagnose EMCI, LMCI, AD and CN patients using a few commonly available neurocognitive tests. The project was successful in creating an accurate, clinically translatable risk-stratified scoring aid for primary care providers to diagnose AD in a fast and inexpensive manner.

## Introduction

Dementia is a neurodegenerative disease characterized by progressive memory loss as a result of neuronal cell death. More than 47 million people worldwide live with dementia and by 2050 that number is expected to increase to 131 million [1]. The most common type of dementia is Alzheimer’s Disease (AD) and Mild Cognitive Impairment (MCI) is often seen as risk state of progression to AD. The latter can be subdivided into early mild cognitive impairment (EMCI) and late mild cognitive impairment (LMCI), as defined in the Alzheimer’s Disease Neuroimaging Initiative (ADNI) database [2]. While there is no cure for dementia, early diagnosis may enable lifestyle changes (such as diet and exercise), neurocognitive enrichment, and therapeutic treatment that may temporarily improve symptoms or slow the rate of decline of symptoms, thereby improving the quality of life [3].

The core clinical criteria for the diagnosis of MCI and AD are neuropsychological tests [4,5]. Fluid and imaging biomarker tests, such as cerebrospinal fluid markers and p-tau, may in some cases supplement standard clinical tests in specialized clinical settings [6]. A large array of neurocognitive tests are currently used to detect cognitive impairment and classify amongst normal controls (CN), EMCI, LMCI and AD [7-8]. Many studies have identified a few top classifiers using logistic regression and machine learning methods [9–18]. Some studies have also used MRI and genetic data in conjunction with neurocognitive measures for classification [19-20]. However, most of these studies to date performed binary classification (i.e., between CN and AD or CN and MCI) [17,21]. Classifying CN, EMCI, LMCI and AD remains challenging.

Deep learning is increasingly being used in medicine, including classification of diseases to aid diagnosis [22-24]. Deep learning, or machine learning in general, uses algorithms to learn the relationship amongst different data elements to inform outcomes. In contrast to traditional analysis methods (such as logistic regression), the specific relationships amongst different input variables with outcome variables do not need to be explicitly specified a priori. Neural networks, for example, are made up of a collection of connected nodes that model the neurons present in a human brain [25]. Each connection, like the synapses in a brain, transmits and receives signals to other nodes. Each node and the connections it forms are initialized with weights which are adjusted throughout training and create mathematical relationships between the input data and the outcomes. Deep learning is well-suited to analyze complex and large datasets where input and output variables cannot be readily parameterized.

The goal of this study was to compare different feature-selection algorithms and develop a deep-learning algorithm to identify the top neurocognitive test scores that accurately classify normal control, early MCI, late MCI and Alzheimer’s disease. From these findings, we further constructed a novel simplified risk score model to classify normal, EMCI, LMCI and AD for clinical use.

## Methods

### Study Population

Data used in this study was obtained from the Alzheimer’s Disease Neuroimaging Initiative (ADNI) database (adni.loni.usc.edu). Patients were taken from the ADNI1, ADNIGO, ADNI2, and ADNI3 patient sets. **Figure 1** shows the flowchart for patient selection. The inclusion criteria were a confirmed diagnosis from screening to the baseline visit and the exclusion criteria were greater than 20% of patient data missing. The total sample size in the study was 1937 patients, with 1743 being randomly assigned to the training dataset and 194 being assigned to the testing dataset before any feature selection or feature engineering was performed. The dataset was the ADNI database, which is a multi-institutional data source, which had built in independent data sets. Future studies will further test using additional independent datasets. Of the 1937 participants that met the inclusion criteria, 516 patients were diagnosed as CN, 383 were diagnosed as EMCI, 644 were diagnosed as LMCI, and 394 were diagnosed as AD.

**Figure 1.**
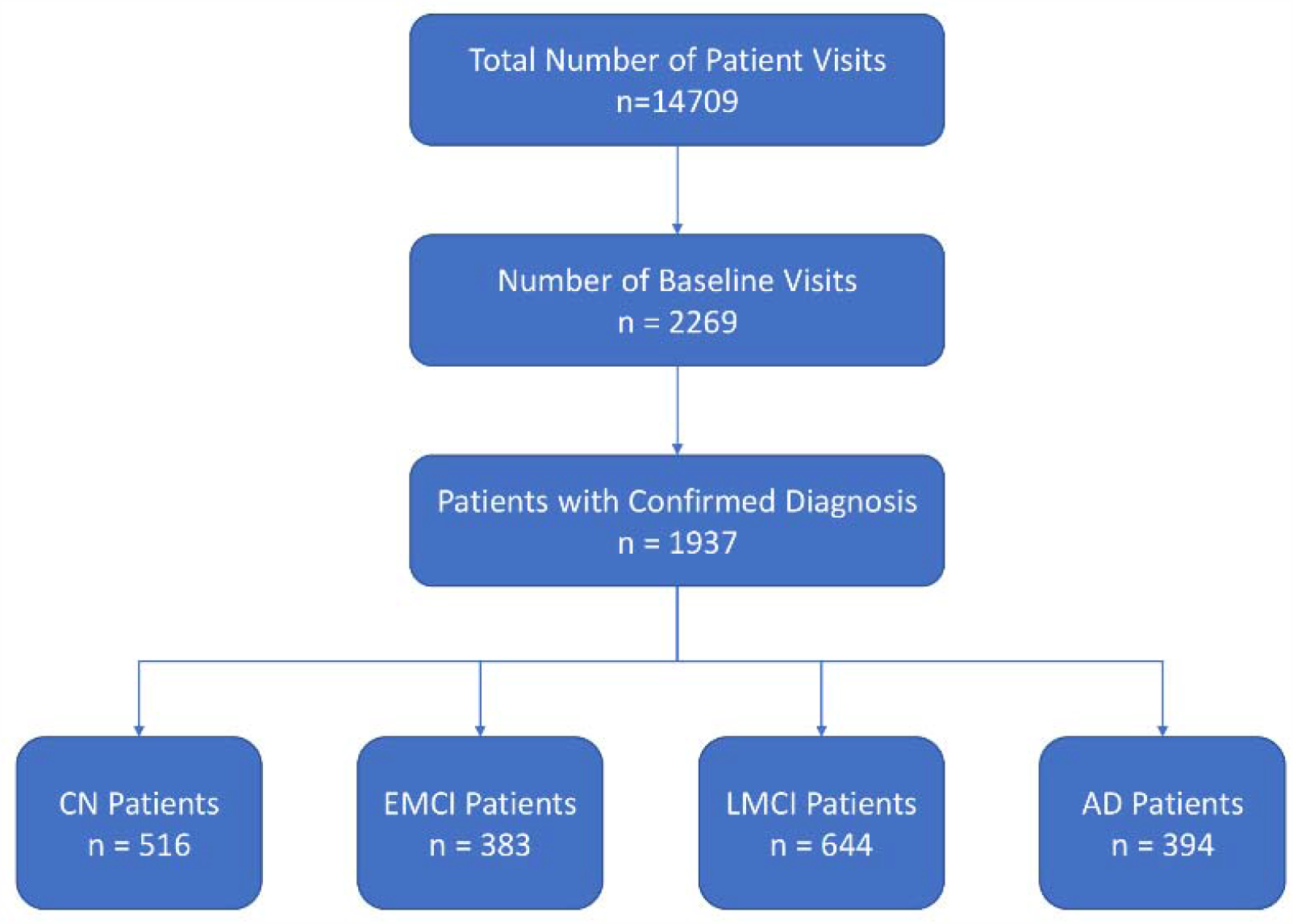
Flowchart of patient selection.

### Data Preprocessing

We evaluated about 100 input variables (i.e., test scores, demographic information, and biomarkers). Correlation matrix analysis showed that 47 variables had a correlation coefficient above 0.5 and were determined to be correlated, which merited exclusion from further analysis. In addition, 29 variables were missing in >20% of patients and they were also excluded from analysis. For the rest of the variables, missing data (most of which had < 10% data missing) was imputed with Classification and Regression Trees (CART) using Multivariate Imputation by Chained Equations (MICE) in R, a statistical analysis software (version 4.0.0) [26]. Although regional volumes were available through the FreeSurfer pipeline, >30% were missing and regional volumes were thus not included in the analysis. Intracranial volume was included with imputation because <20% was missing.

The following neurocognitive tests, demographics, comorbidities and other variables were used in our analysis. The neuropsychological scores included ADAS11 (Unweighted sum of 11 items from The Alzheimer’s Disease Assessment Scale-Cognitive Subscale (ADAS-Cog)), ADAS13 (Unweighted sum of 13 items from ADAS-Cog), ADASQ4 (Score from Task 4 (Word Recognition) of the Alzheimer’s Disease Assessment Scale (ADAS)), CDRSB, FAQ (Functional Activities Questionnaire), LDELTOTAL, MMSE, RAVLT_forgetting (Rey’s Auditory Verbal Learning Test – Forgetting score), RAVLT_immediate (Rey’s Auditory Verbal Learning Test – Immediate Recall score), RAVLT_learning (Rey’s Auditory Verbal Learning Test – Learning Score), RAVLT_percentage_forgetting (Rey’s Auditory Verbal Learning Test – Percent Forgetting), TRABSCOR (Trail Making Test Part B Time), mPACCdigit, and mPACCtrailsB. The extracted imaging parameters that were used included intracranial volume (ICV), volume of ventricles, and whole brain volume. APOE4 status was also included. The demographics included age, sex, race, ethnicity, education level (PTEDUCAT). The outcomes were 4 diagnosis classes: AD, LMCI, EMCI, and CN based on comprehensive clinical diagnosis as provided in the dataset.

### Neural Network Model

Ranking of feature importance was first conducted among cognitive tests, demographic information, genetic tests, and extracted biomarkers. Five different feature selection methods were utilized to identify the most predictive variables: Information Gain, Boruta Random Forest, Recursive Feature Elimination with the Random Forest Classifier, Logistic Regression with LASSO/L1 regularization, and Permutation Importance in Keras. The scikit-learn library was used for Recursive Feature Elimination and Logistic Regression analyses. The Boruta package in R for Boruta Random Forest and Weka for Information Gain. To conduct Permutation Importance analysis, a separate neural network was trained with all features available rather than just the top few variables [27]. This network consisted of 5 layers, a BatchNormalization layer followed by 2 fully connected (FC) dense layers followed by a dropout layer and finally another fully connected dense layer. The first FC layer consisted of 24 neurons with the ReLU activation function and the second FC layer consisted of 16 neurons with the ReLU activation function. The dropout layer had a dropout rate of 0.20. The last layer consisted of 4 neurons with the Softmax activation function for multiclass classification. The model was compiled with categorical cross entropy loss with the ADAM optimizer and a learning rate of 0.001 [28]. The top predictors were those that demonstrated statistical significance.

For the deep learning model, a Multi-layer Perceptron (MLP) neural network was constructed with 2 fully connected dense layers for classification followed by a dropout layer and finally a fully connected dense layer. The first two FC layers contained 8 neurons along with the ReLU activation function. The next layer was a dropout layer with dropout rate of 0.15. The last layer contained 4 neurons and used the Softmax activation function for multiclass classification. The model was compiled with categorical cross entropy loss with the ADAM optimizer and a learning rate of 0.003. Additionally, while testing the classification accuracy of the variables selected by Permutation Importance analysis, a BatchNormalization layer was added as the first layer of the network. The top predictors extracted from the global feature selection analysis were used as input for the neural network and the output was the diagnosis class. The dataset was split into 90% training data and 10% testing data using 10-fold cross validation while training the neural network. Diagnosis results were categorized by multiclass classification.

### Risk Score Model Development

A simplified risk score model was constructed using the top 5 global variables (ca. cognitive test scores) identified by the different feature selection methods as followed: 1) For each variable, scores were plotted for the 4 classes of diagnosis and cutoff points were chosen to maximize separation amongst the 4 classes. 2) The cutoff points were then used to construct a point value system for each cognitive test’s score range. This was done by fitting the top cognitive tests in the training dataset against the diagnosis outcome using a Generalized Linear Model (GLM) [29]. 3) The GLM then assigned risk score points for each of the cognitive tests score ranges. A higher number of points for a given score range means that the patient is more likely to have AD. 4) A composite risk score from the sum of the top 5 variables’ risk scores was constructed for each patient. 5) The risk score model was then tested on an independent testing dataset and evaluated using ROC analysis. Risk scores of the testing dataset were plotted for the 4 classes with interpolation smoothing.

We chose only the top five variables because: i) they are manageable for creating the risk score, ii) feature importance dropped significantly after the first five features for multiple machine-learning methods, which provided further validation for the selection of features and avoid potential bias, and iii) limiting to a few features (instead of all features) prevents overfitting in training the neural network and the risk score models.

### Performance Evaluation and Statistical Analysis

Statistical analyses were performed using SPSS v26.Frequencies and percentages for categorical variables between the stages of AD were compared in a pair-wise fashion using χ^2^ tests. Continuous variables, which were denoted as median (IQR), were first tested for normality with the Lilliefors corrected Kolmogorov-Smirnov test. If they were shown to not have a normal distribution, further comparison was done in a pair-wise fashion between groups using the nonparametric Kruskal-Wallis test. P-values < 0.05 were considered statistically significant.

ROC analysis was used to evaluate the performance of the NN and the risk score model, in which training data was first split into 90% for training and 10% for testing using 10-fold cross validation and then tested on an independent testing set. The AUC calculation was binary, in which one class was contrasted with the rest of the classes (one versus rest) and this was repeated for each of the 4 classes. The sensitivity and specificity reported were taken as an average of the binary sensitivity and specificity of each class. The 95% Confidence Interval (CI) for the AUC was obtained through bootstrapping the neural network’s predictions 1000 times.

## RESULTS

**Table 1** shows the demographic data for CN (n=516), EMCI (n=383), LMCI (n=644), and AD (n=394) groups. Age was not significantly different between groups except between CN and LMCI and between the LMCI and AD groups. Race and ethnicity did not differ significantly between groups. The median education level did not differ significantly between any pair except between AD versus the other classes.

**Table 1.**
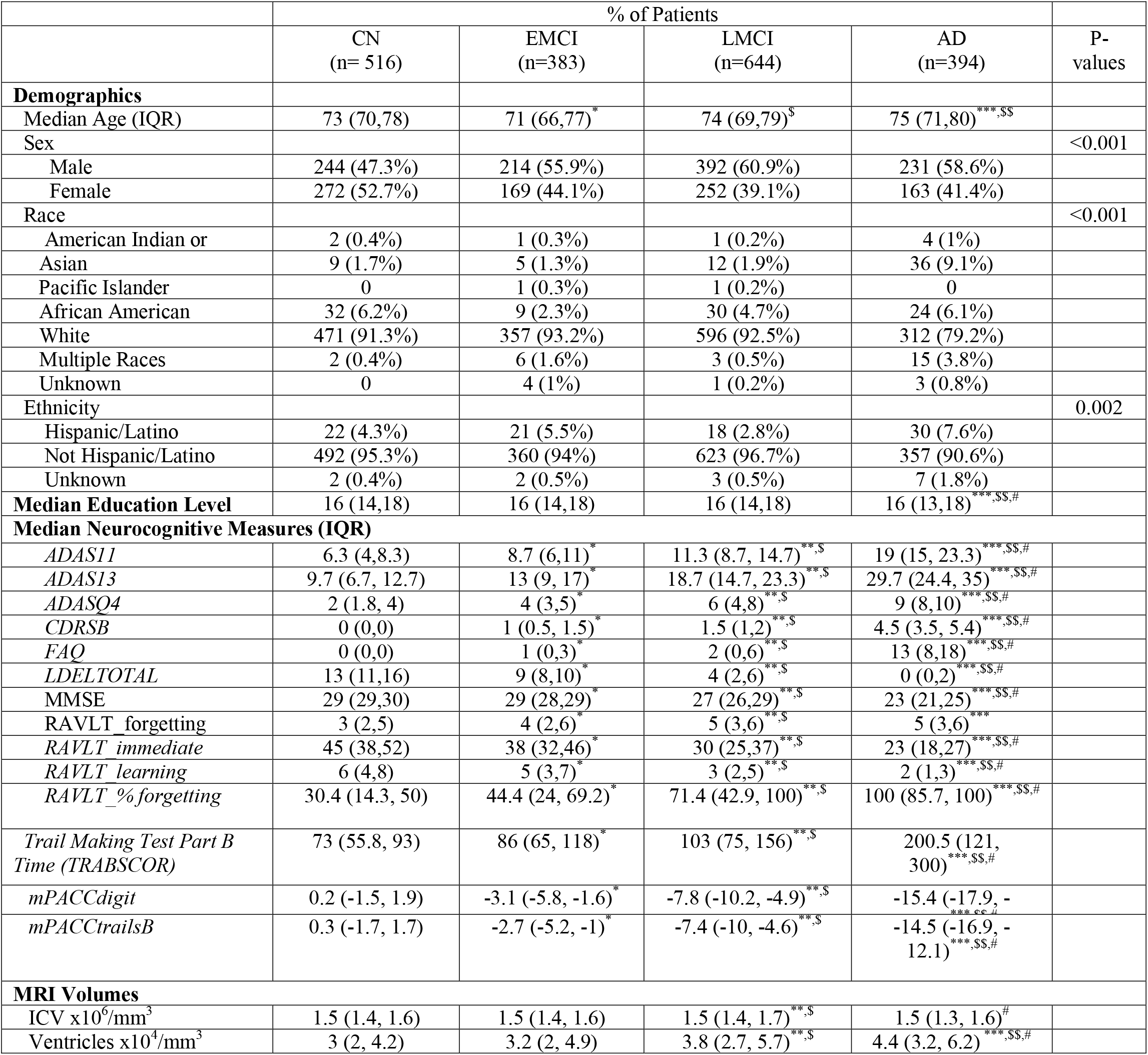
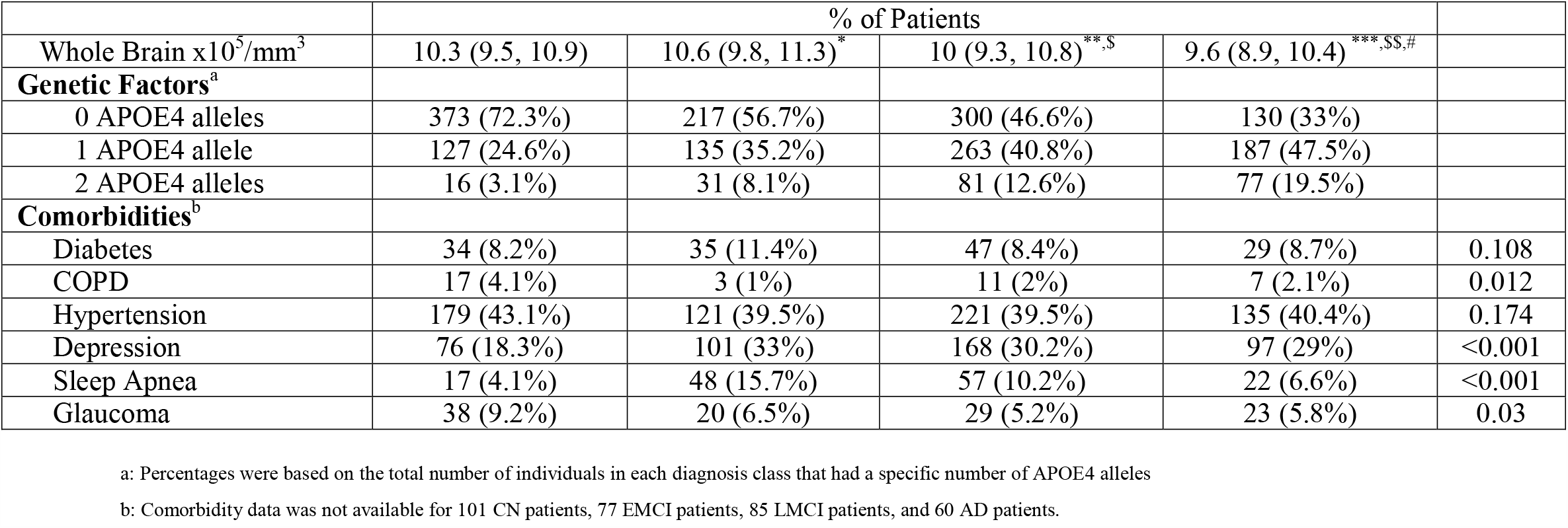
Demographic Information, Neurocognitive Tests, MRI-extracted biomarkers, and Genetic Factors among CN, EMCI, LMCI, AD. Continuous variables are expressed as median (IQR) and the pairwise Kruskal Wallis test is employed. The χ^2^ test is used to identify significance between classes of categorical variables. P-values displayed are with Bonferroni’s correction. Pairwise comparisons are represented by symbols, where a * indicates a statistical difference between the CN and EMCI groups, a ** indicates a statistical difference between the CN and LMCI groups, a *** indicates a statistical difference between the CN and AD groups, a $ indicates a statistical difference between the EMCI and LMCI groups, a $$ indicates a statistical difference between the EMCI and AD groups, and a # indicates a statistical difference between the LMCI and AD groups.

|  | % of Patients |  |  |  | P-values |
| --- | --- | --- | --- | --- | --- |
|  | CN<br>(n= 516) | EMCI<br>(n=383) | LMCI<br>(n=644) | AD<br>(n=394) |  |
| **Demographics** |  |  |  |  |  |
| Median Age (IQR) | 73 (70,78) | 71 (66,77)* | 74 (69,79)\$ | 75 (71,80)***,\$\$ | |
| Sex |  |  |  |  | <0.001 |
| Male | 244 (47.3%) | 214 (55.9%) | 392 (60.9%) | 231 (58.6%) |  |
| Female | 272 (52.7%) | 169 (44.1%) | 252 (39.1%) | 163 (41.4%) |  |
| Race |  |  |  |  | <0.001 |
| American Indian or | 2 (0.4%) | 1 (0.3%) | 1 (0.2%) | 4 (1%) |  |
| Asian | 9 (1.7%) | 5 (1.3%) | 12 (1.9%) | 36 (9.1%) |  |
| Pacific Islander | 0 | 1 (0.3%) | 1 (0.2%) | 0 |  |
| African American | 32 (6.2%) | 9 (2.3%) | 30 (4.7%) | 24 (6.1%) |  |
| White | 471 (91.3%) | 357 (93.2%) | 596 (92.5%) | 312 (79.2%) |  |
| Multiple Races | 2 (0.4%) | 6 (1.6%) | 3 (0.5%) | 15 (3.8%) |  |
| Unknown | 0 | 4 (1%) | 1 (0.2%) | 3 (0.8%) |  |
| Ethnicity |  |  |  |  | 0.002 |
| Hispanic/Latino | 22 (4.3%) | 21 (5.5%) | 18 (2.8%) | 30 (7.6%) |  |
| Not Hispanic/Latino | 492 (95.3%) | 360 (94%) | 623 (96.7%) | 357 (90.6%) |  |
| Unknown | 2 (0.4%) | 2 (0.5%) | 3 (0.5%) | 7 (1.8%) |  |
| **Median Education Level** | 16 (14,18) | 16 (14,18) | 16 (14,18) | 16 (13,18)***,\$\$,# | |
| **Median Neurocognitive Measures (IQR)** |  |  |  |  |  |
| *ADAS11* | 6.3 (4,8.3) | 8.7 (6,11)* | 11.3 (8.7, 14.7)**,\$ | 19 (15, 23.3)***,\$\$,# | |
| *ADAS13* | 9.7 (6.7, 12.7) | 13 (9, 17)* | 18.7 (14.7, 23.3)**,\$ | 29.7 (24.4, 35)***,\$\$,# | |
| *ADASQ4* | 2 (1.8, 4) | 4 (3,5)* | 6 (4,8)**,\$ | 9 (8,10)***,\$\$,# | |
| *CDRSB* | 0 (0,0) | 1 (0.5, 1.5)* | 1.5 (1,2)**,\$ | 4.5 (3.5, 5.4)***,\$\$,# | |
| *FAQ* | 0 (0,0) | 1 (0,3)* | 2 (0,6)**,\$ | 13 (8,18)***,\$\$,# | |
| *LDELTOTAL* | 13 (11,16) | 9 (8,10)* | 4 (2,6)**,\$ | 0 (0,2)***,\$\$,# | |
| *MMSE* | 29 (29,30) | 29 (28,29)* | 27 (26,29)**,\$ | 23 (21,25)***,\$\$,# | |
| *RAVLT_forgetting* | 3 (2,5) | 4 (2,6)* | 5 (3,6)**,\$ | 5 (3,6)*** | |
| *RAVLT_immediate* | 45 (38,52) | 38 (32,46)* | 30 (25,37)**,\$ | 23 (18,27)***,\$\$,# | |
| *RAVLT_learning* | 6 (4,8) | 5 (3,7)* | 3 (2,5)**,\$ | 2 (1,3)***,\$\$,# | |
| *RAVLT_%forgetting* | 30.4 (14.3, 50) | 44.4 (24, 69.2)* | 71.4 (42.9, 100)**,\$ | 100 (85.7, 100)***,\$\$,# | |
| *Trail Making Test Part B Time (TRABSCOR)* | 73 (55.8, 93) | 86 (65, 118)* | 103 (75, 156)**,\$ | 200.5 (121, 300)***,\$\$,# | |
| *mPACCdigit* | 0.2 (-1.5, 1.9) | -3.1 (-5.8, -1.6)* | -7.8 (-10.2, -4.9)**,\$ | -15.4 (-17.9, -12.1)***,\$\$,# | |
| *mPACCtrailsB* | 0.3 (-1.7, 1.7) | -2.7 (-5.2, -1)* | -7.4 (-10, -4.6)**,\$ | -14.5 (-16.9, -12.1)***,\$\$,# | |
| **MRI Volumes** |  |  |  |  |  |
| ICV x10<sup>6</sup>/mm<sup>3</sup> | 1.5 (1.4, 1.6) | 1.5 (1.4, 1.6) | 1.5 (1.4, 1.7)**,\$ | 1.5 (1.3, 1.6)# | |
| Ventricles x10<sup>4</sup>/mm<sup>3</sup> | 3 (2, 4.2) | 3.2 (2, 4.9) | 3.8 (2.7, 5.7)**,\$ | 4.4 (3.2, 6.2)***,\$\$,# | |

|  | % of Patients |  |  |  |  |
| --- | --- | --- | --- | --- | --- |
| Whole Brain $\times 10^5/\text{mm}^3$ | 10.3 (9.5, 10.9) | 10.6 (9.8, 11.3)<sup>*</sup> | 10 (9.3, 10.8)<sup>**,\$</sup> | 9.6 (8.9, 10.4)<sup>***,\$\$,#</sup> | |
| **Genetic Factors<sup>a</sup>** |  |  |  |  |  |
| 0 APOE4 alleles | 373 (72.3%) | 217 (56.7%) | 300 (46.6%) | 130 (33%) |  |
| 1 APOE4 allele | 127 (24.6%) | 135 (35.2%) | 263 (40.8%) | 187 (47.5%) |  |
| 2 APOE4 alleles | 16 (3.1%) | 31 (8.1%) | 81 (12.6%) | 77 (19.5%) |  |
| **Comorbidities<sup>b</sup>** |  |  |  |  |  |
| Diabetes | 34 (8.2%) | 35 (11.4%) | 47 (8.4%) | 29 (8.7%) | 0.108 |
| COPD | 17 (4.1%) | 3 (1%) | 11 (2%) | 7 (2.1%) | 0.012 |
| Hypertension | 179 (43.1%) | 121 (39.5%) | 221 (39.5%) | 135 (40.4%) | 0.174 |
| Depression | 76 (18.3%) | 101 (33%) | 168 (30.2%) | 97 (29%) | <0.001 |
| Sleep Apnea | 17 (4.1%) | 48 (15.7%) | 57 (10.2%) | 22 (6.6%) | <0.001 |
| Glaucoma | 38 (9.2%) | 20 (6.5%) | 29 (5.2%) | 23 (5.8%) | 0.03 |

a: Percentages were based on the total number of individuals in each diagnosis class that had a specific number of APOE4 alleles
b: Comorbidity data was not available for 101 CN patients, 77 EMCI patients, 85 LMCI patients, and 60 AD patients.

With few exceptions, all neurocognitive test scores and mPACC tests showed pairwise differences between groups. The MRI-extracted parameters were significant between CN and AD and between CN and LMCI, but not significant between the other pairwise comparison. APOE4 was significant different in all pairwise comparisons. Sleep apnea and depression were the only significant comorbidities between groups.

**Figure 2** shows the results of the rankings by importance from the 5 feature selection methods performed on the training dataset and **Table 2** lists the top 9 features. CDRSB was the most frequently identified top feature amongst the top 5 feature selection methods (5 out of 5), followed by LDELTOTAL (4 out of 5), mPACCdigit (4 out of 5), mPACCtrailsB (3 out of 5), MMSE (2 out of 5).

**Table 2.** Top 9 clinical variables ranked by 5 feature selection methods

| Ranking | Information Gain | Boruta Random Forest | Recursive Feature Elimination | Logistic Regression with LASSO regularization | Permutation Importance |
| --- | --- | --- | --- | --- | --- |
| 1 | CDRSB | CDRSB | CDRSB | CDRSB | LDELTOTAL |
| 2 | LDELTOTAL | LDELTOTAL | LDELTOTAL | APOE4 | CDRSB |
| 3 | mPACCdigit | mPACCtrailsB | mPACCdigit | FAQ | MMSE |
| 4 | mPACCtrailsB | mPACCdigit | mPACCtrailsB | PTEDUCAT | mPACCdigit |
| 5 | ADAS13 | MMSE | PTEDUCAT | ADAS13 | WholeBrain |
| 6 | FAQ | FAQ | MMSE | RAVLT_immediate | ICV |
| 7 | MMSE | PTEDUCAT | ADAS13 | RAVLT_forgetting | PTEDUCAT |
| 8 | ADAS11 | ADAS13 | FAQ | Ventricles | RAVLT_perc_forgetting |
| 9 | ADASQ4 | ADAS11 | AGE | LDELTOTAL | ICV |

ADAS11: Unweighted sum of 11 items from The Alzheimer's Disease Assessment Scale-Cognitive Subscale (ADAS-Cog)
ADAS13: Unweighted sum of 13 items from ADAS-Cog
ADASQ4: Score from Task 4 (Word Recognition) of the Alzheimer's Disease Assessment Scale (ADAS)
CDRSB: Clinical Dementia Rating - Sum of Boxes Score
FAQ: Functional Activities Questionnaire
ICV: Intracranial Volume
LDELTOTAL: Delayed Total Recall
MMSE: Mini-Mental State Examination
PTEDUCAT: Education Level
RAVLT\_forgetting: Rey's Auditory Verbal Learning Test – Forgetting score
RAVLT\_immediate: Rey's Auditory Verbal Learning Test – Immediate Recall score
RAVLT\_percentage\_forgetting: Rey's Auditory Verbal Learning Test – Percent Forgetting
mPACCdigit: Modified Preclinical Alzheimer Cognitive Composite with Digit test
mPACCtrailsB: Modified Preclinical Alzheimer Cognitive Composite with Trails test

**Figure 2.**
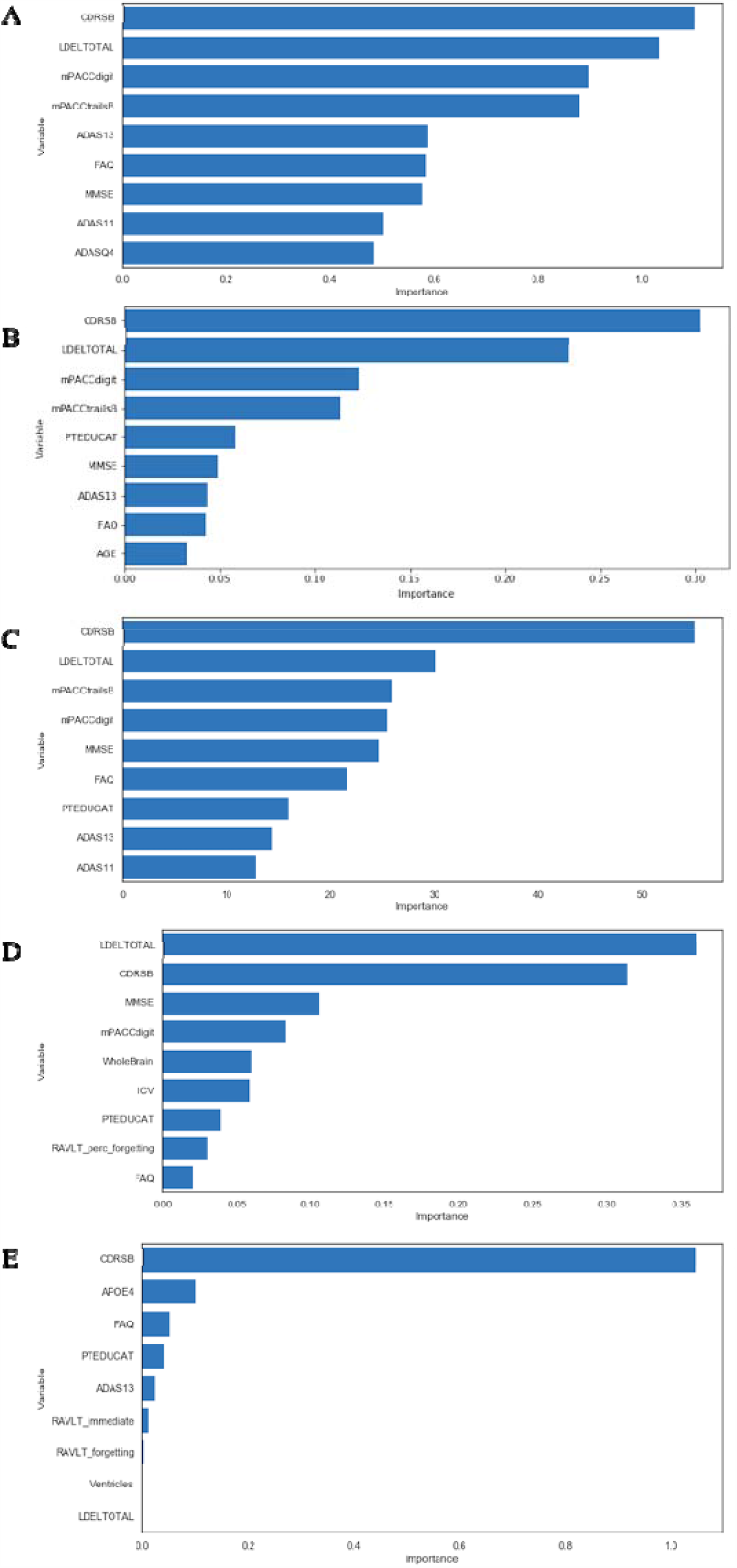
Feature ranking for (A) Information Gain, (B) Recursive Feature Elimination with the Random Forest classifier, (C) Boruta Random Forest, (D) Permutation Importance, and (E) Logistic Regression with LASSO/L1 regularization

### Neural Network Model for Classification

Classification was performed using the top 5 features. The performances on the testing data for the 5 methods are summarized in **Table 3**. AUCs for the Information Gain, Boruta Random Forest, Recursive Feature Elimination with the Random Forest Classifier, Logistic Regression with LASSO/L1 regularization, and Permutation Importance were 0.978, 0.984, 0.983, 0.906 and 0.982, respectively, on the testing dataset. The classifier selected by Boruta Random Forest performed the best in terms of AUC, but the classifier selected by Recursive Feature Elimination performed better in terms of accuracy, sensitivity, and specificity. By comparison, classification using CDRSB, LDELTOTAL, mPACCdigit, mPACCtrailsB, and MMSE individually yielded an AUC of 0.8899, 0.8957, 0.8619, 0.8624, and 0.7808 respectively, on the testing dataset.

**Table 3.** Performance of the variables selected from the 5 feature selection methods in our NN. Bracket values indicate 95% confident intervals.

|  | Boruta<br>Random<br>Forest | Recursive<br>Feature<br>Elimination | Permutation<br>Importance | Information<br>Gain | Logistic<br>Regression |
| --- | --- | --- | --- | --- | --- |
| AUC | 0.981<br>[0.971-0.99] | 0.986<br>[0.978,0.994] | 0.982<br>[0.972,0.99] | 0.978<br>[0.966,0.988] | 0.910<br>[0.887, 0.932] |
| Accuracy (%) | 91.24 | 90.72 | 90.72 | 90.21 | 73.71 |
| Sensitivity (%) | 90.88 | 90.47 | 90.30 | 89.64 | 73.09 |
| Specificity (%) | 96.94 | 96.78 | 96.76 | 96.53 | 90.75 |

### Risk Score Model

We then developed a simplified risk score model using the same top 5 variables from our deep-learning analysis. **Figure 3** shows an example of the CDRSB cognitive test scores for 4 different classes. The cutoff points that maximized separation between the 4 classes were 0.1, 1.3, and 3.5, between CN and EMCI, between EMCI and LMCI, and between LMCI and AD group, respectively. The point values for individual test score ranges are summarized in **Table 4**. The composite score from the top 5 cognitive tests (CDRSB, LDELTOTAL, mPACCdigit, mPACCtrailsB, and MMSE) were constructed using a GLM. The classification results on the testing dataset are shown in **Figure 4**. The risk score system classified the four groups accurately. The performance of the risk score model yielded an AUC of 0.963 [95% CI: 0.945-0.975], sensitivity of 88.06% and specificity of 96.16%, and accuracy of 89.18% for the testing dataset.

**Table 4:**
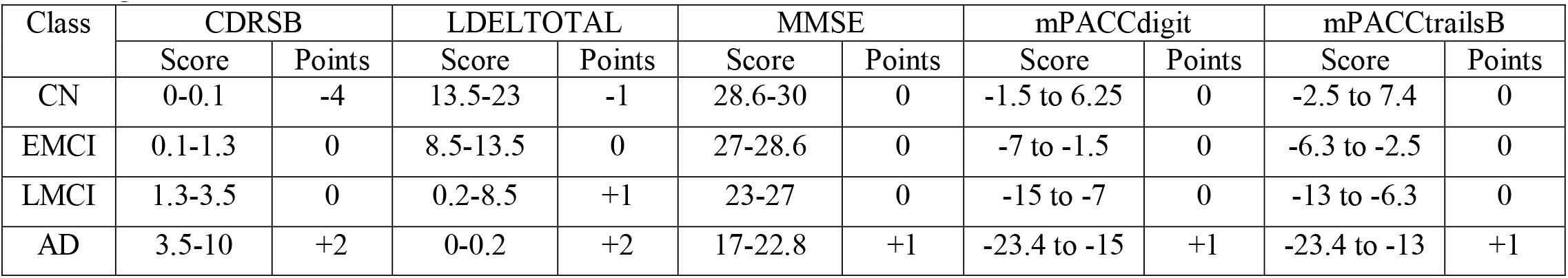
Points given by the risk score model for each cognitive test and per diagnosis class on the training dataset.

| Class | CDRSB |  | LDELTOTAL |  | MMSE |  | mPACCdigit |  | mPACCtrailsB |  |
| --- | --- | --- | --- | --- | --- | --- | --- | --- | --- | --- |
|  | Score | Points | Score | Points | Score | Points | Score | Points | Score | Points |
| CN | 0-0.1 | -4 | 13.5-23 | -1 | 28.6-30 | 0 | -1.5 to 6.25 | 0 | -2.5 to 7.4 | 0 |
| EMCI | 0.1-1.3 | 0 | 8.5-13.5 | 0 | 27-28.6 | 0 | -7 to -1.5 | 0 | -6.3 to -2.5 | 0 |
| LMCI | 1.3-3.5 | 0 | 0.2-8.5 | +1 | 23-27 | 0 | -15 to -7 | 0 | -13 to -6.3 | 0 |
| AD | 3.5-10 | +2 | 0-0.2 | +2 | 17-22.8 | +1 | -23.4 to -15 | +1 | -23.4 to -13 | +1 |

**Figure 3.**
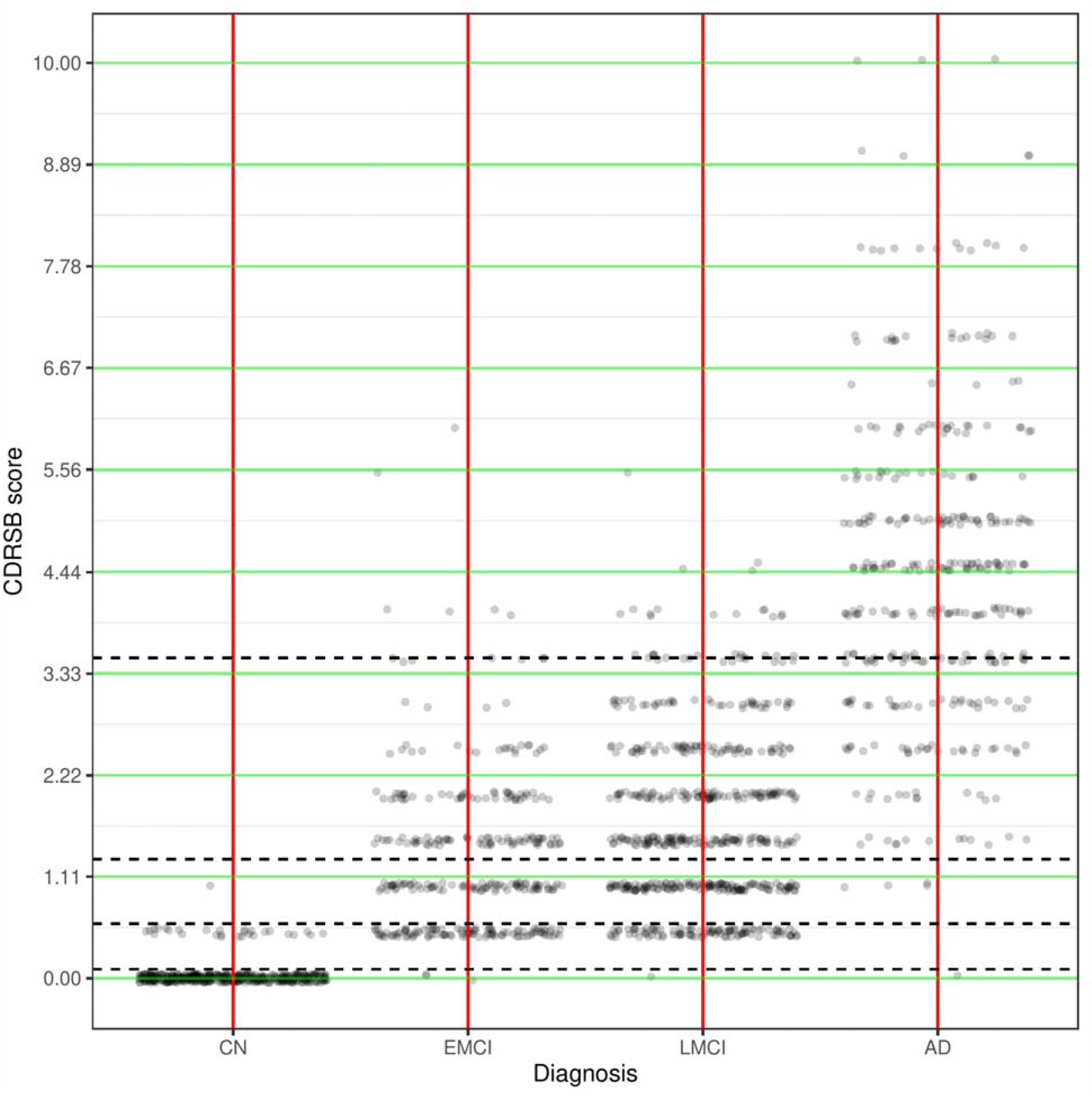
Scatterplot of CDRSB scores versus Patient Diagnosis in the training dataset. The black dashed lines represent the cutoff points that maximize separation between diagnosis classes.

**Figure 4.**
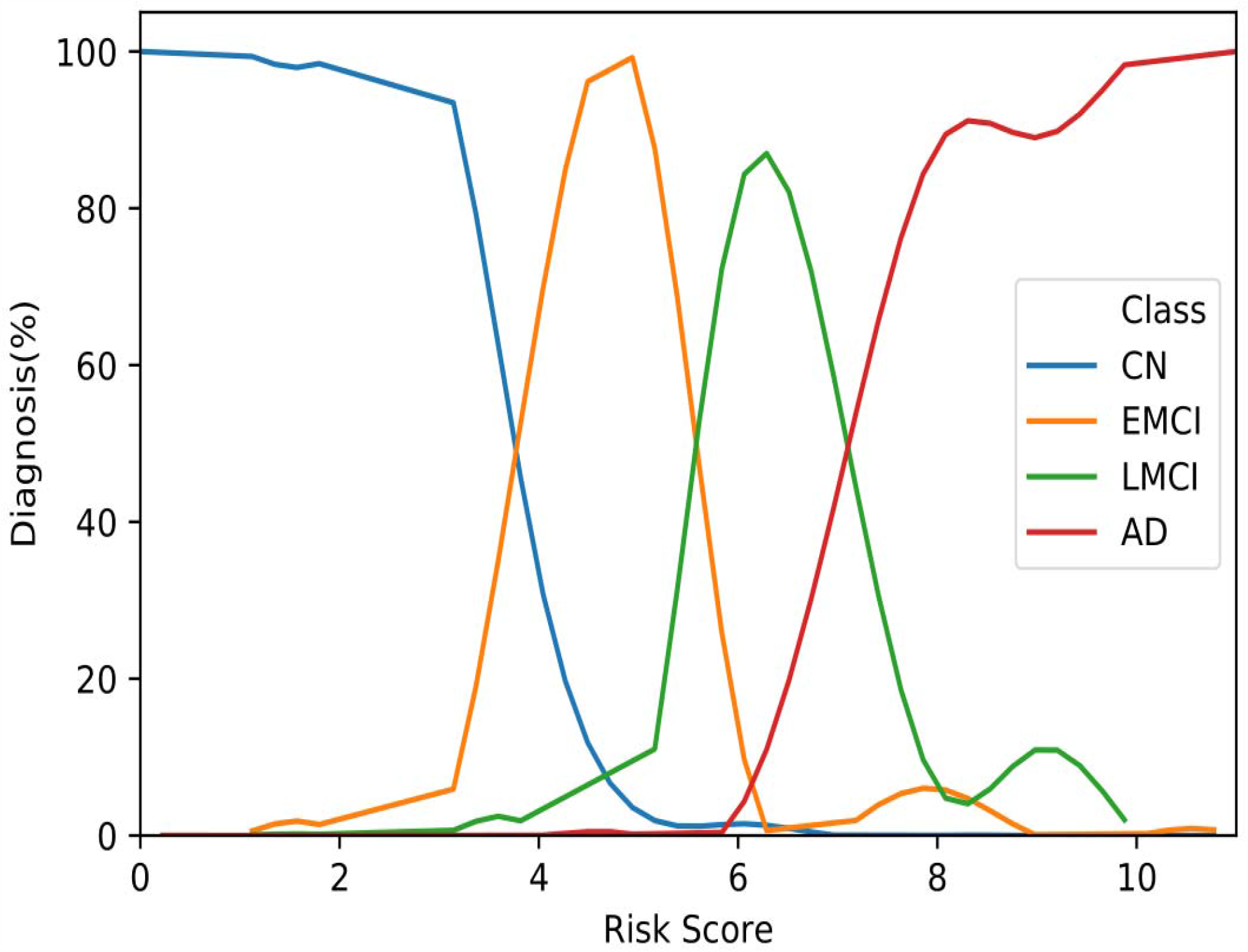
Composite risk score stratification. The scores ranged from 0 to 11, with 11 indicating the greatest risk for developing Alzheimer’s and 0 indicating the lowest.

## Discussion

This study developed a deep-learning algorithm to identify the top neurocognitive test scores that accurately classify normal control, early MCI, late MCI and Alzheimer’s disease. Multiple feature selection methods identified essentially the same set of top variables, providing further corroboration. CDRSB was identified to be a top feature, followed by LDELTOTAL, mPACCdigit, mPACCtrailsB, and MMSE for classification of disease subtypes. Performance indices of the deep-learning model and the simplified score system were highly accurate in classifying the four groups. The best model yielded an AUC of 0.984, and the simplified risk stratification score yielded an AUC of 0.962 for classification on the test dataset. We concluded that only a few neurocognitive tests are needed to accurately classify normal control, early MCI, late MCI and AD.

Clinical diagnosis of AD and MCI involves a large collection of clinical variables. They include two major neurocognitive tests: CDR and MMSE. MMSE and CDR are useful to distinguish between CN and AD but less so amongst EMCI, LMCI and CN (REF). We found that CDR was amongst the top performers to classify amongst CN, EMCI, LMCI, and AD, but MMSE was not.

The ADNI dataset consists of a large array of neurocognitive tests that are not currently being used in clinical settings but could have future applications. With the advances in computing, it becomes possible to use machine learning to analyze the large array of neurocognitive tests to accurately classify CN, EMCI, LMCI, and AD.

Although CDR and MMSE were used in the clinical diagnosis, there are other variables that were highly ranked, thereby providing insights into specific domain of cognitive dysfunction. It is possible that high performance was dominated by a few variables of the same cognitive test group. However, we used correlation-matrix analysis to remove variables that are highly correlated. Specifically, CDR and MMSE was found to be weakly correlated. This is not surprising as CDR and MMSE measure different dimensions of cognitive function. Our approach selected a small set of top predictors among many that are highly predictive of outcome.

The key findings are: i) our NN model was able to diagnose 4 classes, which is not commonly done, ii) our NN model performance is comparable to literature, iii) the combined top neurocognitive scores performed better in distinguishing CN, EMCI, LMCI, and AD than individual scores. Taken together, our NN model and risk score can ultimately improve classification or diagnosis accuracy because it uses multiparametric data. ML can also incorporate longitudinal multiparametric data to predict disease progression.

The top 5 classifiers were all neurocognitive test scores. The CDRSB is rated along 6 domains of functioning, with each domain being rated on a 5-point scale, and the global CDRSB being a function of the scores from these 6 domains [29]. A higher CDRSB indicates more severe impairment. LDELTOTAL measures episodic memory and performance is measured primarily through the amount of a story that is remembered [30]. A lower LDELTOTAL score indicates more severe impairment. The mPACCdigit test measures working memory by asking the patient to repeat back a sequence of digits of increasing length, until they are not able to. The mPACCtrailsB test determines performance of processing speed with a smaller score indicating more severe impairment. Lastly, the MMSE, one of the most clinically used battery tests, is a 30-question questionnaire that is used to screen for dementia and includes tasks that involve registration, recall, and attention. Individually, these tests all perform well in separating AD from CN individuals, but struggle to diagnose MCI subtypes, Indeed, we found classification using all top 5 variables and the derived risk score system outperformed classification using CDRSB, LDELTOTAL, mPACCdigit, mPACCtrailsB, and MMSE individually.

It is interesting to note that intracranial volume (ICV), volume of ventricles, and whole brain volume by MRI were not highly ranked as classifiers of the 4 classes. Volumetric differences could readily distinguish between CN and AD groups but might not readily differentiate between CN and MCI or between MCI subclasses [31]. Other studies investigating hippocampal volume for classifying dementia subtypes showed promise, but it is still challenging for hippocampal volume to accurately classify between CN and MCI patients (Chincarini et al., 2016). We did not include hippocampal volume because it was not readily available in the dataset.

Many studies have previously examined neurocognitive tests and identified a few top neurocognitive classifiers using non-machine learning methods, but they are not discussed here [9,11-13] (see reviews by *Abd Razak et al*. and *De Roeck et al*.) By comparison, only few studies utilized (mostly supervised learning) machine learning methods and some of these studies combined neurocognitive tests and MRI regional brain volumes as input variables [10,14-18] (**Table 5**). *So et al*. used a two-stage approach to classification with the first stage identifying the most important subsections of the MMSE and the second stage used subsections of the Consortium to Establish a Registry for Alzheimer’s Disease (CERAD) assessment [14]. They achieved up to 97% classification accuracy in Stage 1 with an MLP and 75% classification accuracy in Stage 2 with support vector machine. *Lins et al*. investigated a Brazilian dataset and utilized gender, age, study time (in years), AD8, MMSE, Clinical Dementia Rating (CDR), and SVFT scores, and two genetic markers (CYP46A1 and APOE4) [15]. They tested the predictive power using the Random Forest, support vector machine, and Stochastic Gradient Boosting classifiers along with an MLP neural network. They achieved a maximum binary classification accuracy between dementia and CN patients of 96% using the CDR and CYP46 features. *Stamate et al*. identified mPACCdigit, mPACCtrailsB, LDELTOTAL as the top classifiers [10]. They combined these scores with PET and MRI data and achieved an AUC of 0.88 for the binary classification of NC versus dementia. *Chiu et al*. developed NMD-12, a 12-question questionnaire that was shortened from the original 45 question questionnaire from the HAICDDS project by the Information Gain algorithm [16]. They showed that this test performed better than the commonly used MMSE and MoCA tests with an AUC of 0.94 for discriminating between CN and MCI patients and 0.97 for MCI and dementia patients. *Zhu et al*. analyzed a Taiwan cohort and ranked the relative importance of neuropsychological tests using Information Gain, Random Forest, and the Relief algorithm [18]. They classified normal, MCI, VMD, and dementia. They selected a few top ranked features, and their optimized algorithm had an accuracy of 0.81 using Relief feature selection followed by classification with MLP method. *Gill et al*. investigated an MRI-based feature and Modified Barthel Index Score (activities of daily living) for binary classification between CN and MCI [17]. They used supervised machine learning and found the AUC to be 0.86.

**Table 5.**
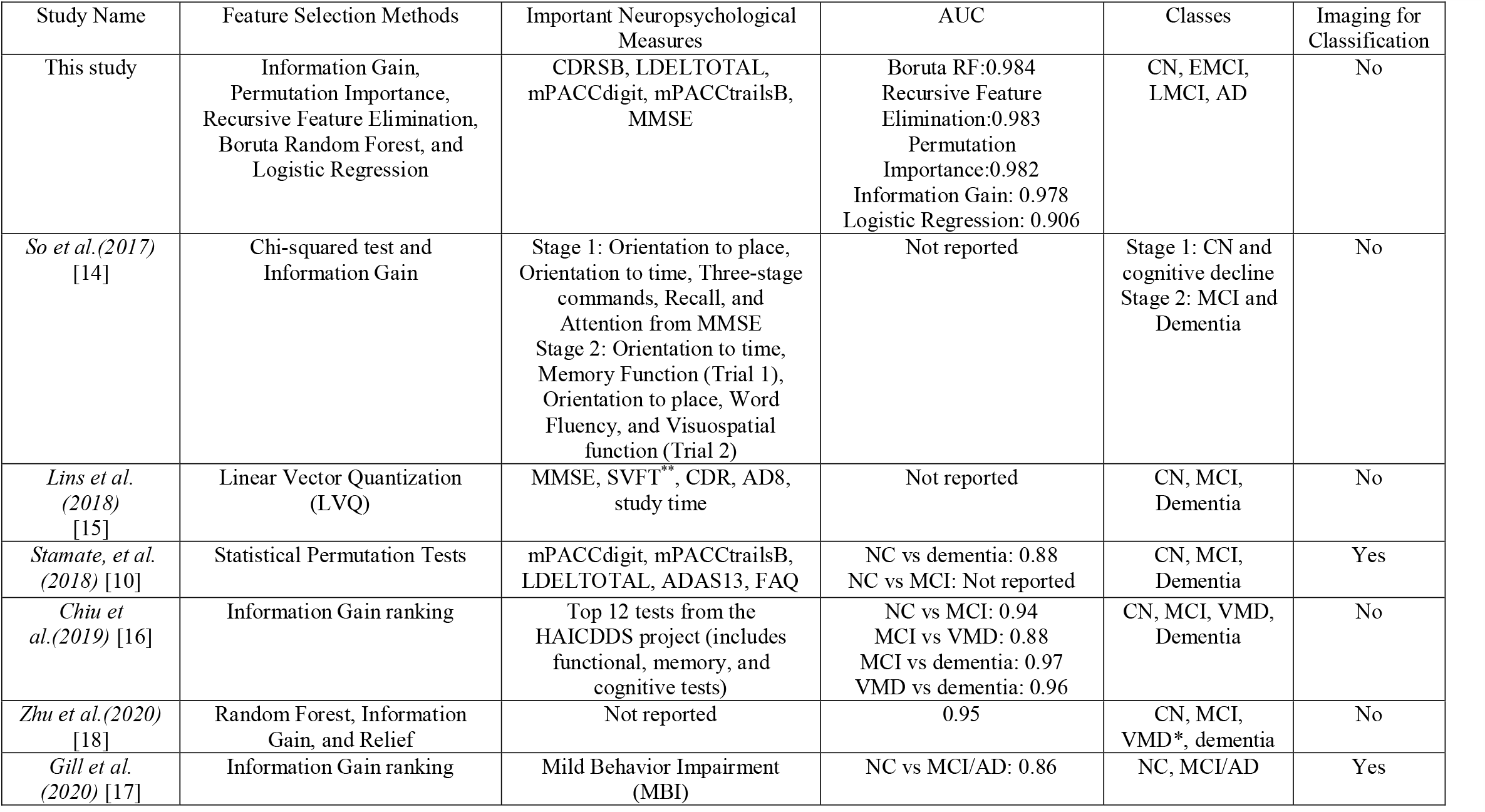
Comparison of machine learning studies in classifying different forms of dementia. VMD: very mild dementia, SVFT: Semantic Verbal Fluency Test, AD8: Dementia Screening Interview

| Study Name | Feature Selection Methods | Important Neuropsychological Measures | AUC | Classes | Imaging for Classification |
| --- | --- | --- | --- | --- | --- |
| This study | Information Gain, Permutation Importance, Recursive Feature Elimination, Boruta Random Forest, and Logistic Regression | CDRSB, LDELTOTAL, mPACCdigit, mPACCtrailsB, MMSE | Boruta RF:0.984<br>Recursive Feature Elimination:0.983<br>Permutation Importance:0.982<br>Information Gain: 0.978<br>Logistic Regression: 0.906 | CN, EMCI, LMCI, AD | No |
| *So et al.(2017)* [14] | Chi-squared test and Information Gain | Stage 1: Orientation to place, Orientation to time, Three-stage commands, Recall, and Attention from MMSE<br>Stage 2: Orientation to time, Memory Function (Trial 1), Orientation to place, Word Fluency, and Visuospatial function (Trial 2) | Not reported | Stage 1: CN and cognitive decline<br>Stage 2: MCI and Dementia | No |
| *Lins et al. (2018)* [15] | Linear Vector Quantization (LVQ) | MMSE, SVFT*, CDR, AD8, study time | Not reported | CN, MCI, Dementia | No |
| *Stamate, et al. (2018)* [10] | Statistical Permutation Tests | mPACCdigit, mPACCtrailsB, LDELTOTAL, ADAS13, FAQ | NC vs dementia: 0.88<br>NC vs MCI: Not reported | CN, MCI, Dementia | Yes |
| *Chiu et al.(2019)* [16] | Information Gain ranking | Top 12 tests from the HAICDDS project (includes functional, memory, and cognitive tests) | NC vs MCI: 0.94<br>MCI vs VMD: 0.88<br>MCI vs dementia: 0.97<br>VMD vs dementia: 0.96 | CN, MCI, VMD, Dementia | No |
| *Zhu et al.(2020)* [18] | Random Forest, Information Gain, and Relief | Not reported | 0.95 | CN, MCI, VMD*, dementia | No |
| *Gill et al. (2020)* [17] | Information Gain ranking | Mild Behavior Impairment (MBI) | NC vs MCI/AD: 0.86 | NC, MCI/AD | Yes |

In sum, our results are comparable or compared favorably with previous studies although comparisons were not made on the same datasets. Our study is novel in that we employed a deep learning method, applied to a large and multi-center ADNI dataset with commonly used measures. We also classified amongst four groups instead of commonly used binary classification in most previous studies (i.e., between normal controls vs AD, or normal controls versus MCI).

This study has several limitations. In this cohort white and non-Hispanic/Latino ethnicities are overrepresented. Some of the comorbidities, such as sleep apnea and depression, showed significant differences between the groups. We also did not include imaging variables. Further evaluation of additional independent datasets, including prospective studies, would improve generalizability of these findings.

An eventual goal of our and other similar approaches is to ultimately create an automated machine learning algorithm and a derived simplified risk score system to help physicians to make more streamlined and accurate diagnoses. Machine learning approaches can help physicians by offering an objective initial assessment and possibly a second opinion of the diagnosis. Moreover, in some other fields of medicine, machine learning can already accurately estimate risk for coronary heart disease [34] and the detection of lung nodules on chest X-rays [35]. In addition to approximating physician skills, machine learning can also detect novel relationships not readily apparent to human perception, especially in large, complex, and longitudinal datasets.

## Conclusion

This study developed a deep-learning algorithm and a simplified risk score to identify the top neurocognitive test scores to classify normal control, early MCI, late MCI and Alzheimer’s disease. We concluded that only a few neurocognitive tests are needed to accurately classify normal control, early MCI, late MCI and AD. Accurate and early diagnosis may lead to better management of the diseases, including interventions that improve symptoms or slow the rate of decline of symptoms.

## Data Availability

All data used in this study is publicly available on the ADNI (Alzheimer's Disease Neuroimaging Initiative website. Due to the public and de-identified nature of the data, an IRB was not needed. This is non-human subject research.

http://adni.loni.usc.edu/

